# Predicting remission after internet-delivered psychotherapy in patients with depression using machine learning and multi-modal data

**DOI:** 10.1101/2021.04.30.21256367

**Authors:** John Wallert, Julia Boberg, Viktor Kaldo, David Mataix-Cols, Oskar Flygare, James J Crowley, Matthew Halvorsen, Fehmi Ben Abdesslem, Magnus Boman, Evelyn Andersson, Nils Hentati Isacsson, Ekaterina Ivanova, Christian Rück

## Abstract

**BACKGROUND:** Whether a patient benefits from psychotherapy or not is arguably a complex process and heterogeneous information extracted from process, genetic, demographic, and clinical data could contribute to the prediction of remission status after psychotherapy. This study applied supervised machine learning with such multi-modal baseline data to predict remission in patients with major depressive disorder (MDD) after completed psychotherapy.

**METHODS:** Eight-hundred ninety-four genotyped adult patients (65.5% women, age range 18-75 years) diagnosed with MDD and treated with guided Internet-based Cognitive Behaviour Therapy (ICBT) at the Internet Psychiatry Clinic in Stockholm were included (2008-2016). Predictor variables from multiple domains were available: demographic, clinical, process (e.g. time to complete online questionnaires), and genetic (polygenic risk scores for depression, education and more). The outcome was remission status post ICBT (cut-off ≤10 on MADRS-S). Data were split into train (60%) and validation (40%) sets based on treatment start date. Predictor selection employed human domain knowledge followed by Recursive Feature Elimination. Model derivation was internally validated through repeated cross-validation resampling. The final random forest model was externally validated against a (i) null, (ii) logit, (iii) XGBoost, and (iv) blended meta-ensemble model on the hold-out validation set. Model transparency was explored through partial dependence and Local Interpretable Model-agnostic Explanations (LIME) analysis.

**RESULTS:** Feature selection retained 45 predictors representing all four predictor types. With unseen validation data, the final random forest model proved reasonably accurate at classifying post ICBT remission (Accuracy 0.656 [0.604, 0.705], P vs null model = 0.004; AUC 0.687 [0.631, 0.743]), slightly better vs logit (bootstrap D =1.730, P = 0.084) but not vs XGBoost (D = 0.463, P = 0.643). Transparency analysis showed model usage of all predictor types at both the group and individual patient level.

**CONCLUSION:** A new, multi-modal classifier for predicting MDD remission status after ICBT treatment in routine psychiatric care was derived and empirically validated. The multi-modal approach to predicting remission may inform tailored treatment, and deserves further investigation.

**One-sentence Summary:** Predicting remission of depression in adults after psychotherapy

## BACKGROUND

Major Depressive Disorder (MDD) is a common mental disorder and a leading cause of disability affecting >260 million individuals worldwide.^1^ In Europe and US alone, around 35 million are estimated to suffer from untreated MDD with a treatment gap of approximately 50%.^2 3^ Treatment of MDD includes psychotropic medication, psychotherapy, and their combination. Improved treatment accessibility for MDD would produce an estimated net benefit of ∼230 billion USD in productivity gains worldwide.^4^ For mild to moderate MDD, cognitive behavioural therapy (CBT) and its more cost-effective online version (ICBT) are empirically supported^3 5^ and recommended by guidelines.^6 7^ However, a substantial portion of individuals – estimates range from 10 to 60% – do not respond sufficiently to ICBT.^8 9^As there is room for improvement, researchers have begun to investigate what variables predict symptom reduction,^3 8 10^ remission status,^11^ and other outcomes proximal to ICBT response such as adherence,^12^ and dropout.^13^ Identifying replicable predictors could inform clinical decision making allowing for better tailored intervention and care for these patients.

Depression is a polygenic condition^14^ which phenotypical expression is also influenced by environmental factors and gene-environment interactions.^15^ Consequently, CBT response is also likely to be a multi-factorial trait for which a range of predictors have been identified, including prior psychological treatment,^16^ baseline symptom severity,^8 10 16^, time-updated weekly symptom severity,^17^ disability status,^10^ quality of life, computer comfortability,^18^ education,^10^ and sex.^8^ Also, process-specific ICBT predictors,^12^ and Polygenic Risk Scores (PRS)^19^ have been suggested. A common characteristic of predictive modelling studies of ICBT outcomes is that prediction performance has room for improvement, suggesting larger sample sizes and a richer multi-modal predictor approach to ICBT outcome prognostication in patients with MDD. To the extent of our knowledge, this has not been tried before. Moreover, the application of machine learning methodology for ICBT outcome prediction has been rare yet may strengthen model derivation and performance.^20 21^ The present study thus investigated a multi-modal data approach to predict post ICBT remission status under ecologically valid conditions using a large sample of patients with MDD treated with ICBT at the national Internet Psychiatry Clinic (IPSY) in Sweden. Critically, all patients had time-stamped online behaviour registered in the online treatment platform and had been genotyped. This enabled inclusion of a wide range of pre-treatment predictors, including demographic and clinical variables, process variables (e.g., how long it took for a patient to complete a baseline questionnaire), and PRSs for different potentially predictive traits (e.g., PRS for MDD, other psychiatric disorders, intelligence). Both well-known log-linear modelling and more flexible non-linear algorithms were applied in a machine learning pipeline of predictor selection and model derivation followed by external validation for predicting post ICBT remission in MDD.

## METHODS

### Ethical considerations

The present study complies with the Declaration of Helsinki and was granted ethics approval by the Regional Ethics Board in Stockholm (dnr 2009/1089-31/2 & 2014/1897-31).

### Patients and setting

Details on patient referral, recruitment, treatment, and study setting are reported elsewhere.^19^ In summary, 894 patients (≥18 yrs) with MDD undergoing a standardized therapist-supported ICBT protocol were recruited from the Internet Psychiatry Clinic (IPSY) in Stockholm from 2008 through 2016. IPSY is a specialized treatment and research unit and the largest ICBT unit in Sweden. Participants donated blood for genotyping and much of their ICBT process activity on the online platform was logged (e.g., time of day and duration of questionnaire completion). The procedure encompassed online screening, on-site psychiatric assessment, initial exclusion/referral (if unable to read or write Swedish, severe MDD, moderate/high suicide risk, recent medication changes, bipolar or psychotic disorder, drug dependence). Treatment with ICBT was for 12 weeks in sequential homework assignment format, with first follow-up at post measurement at treatment completion and second follow-up 3-6 months post treatment start.^22^

### Outcome

The Montgomery-Åsberg Depression Rating Scale-Self report (MADRS-S) completed at post treatment classifying a total score ≤10 as remission, and >10 as no remission. The MADRS-S is a widely used instrument for assessing MDD^23 24^ that is validated for the present study cut-off.^25^

### Modelling

Modelling involved several steps. See **Figure 1** for a modelling flowchart.

**Figure 1.**
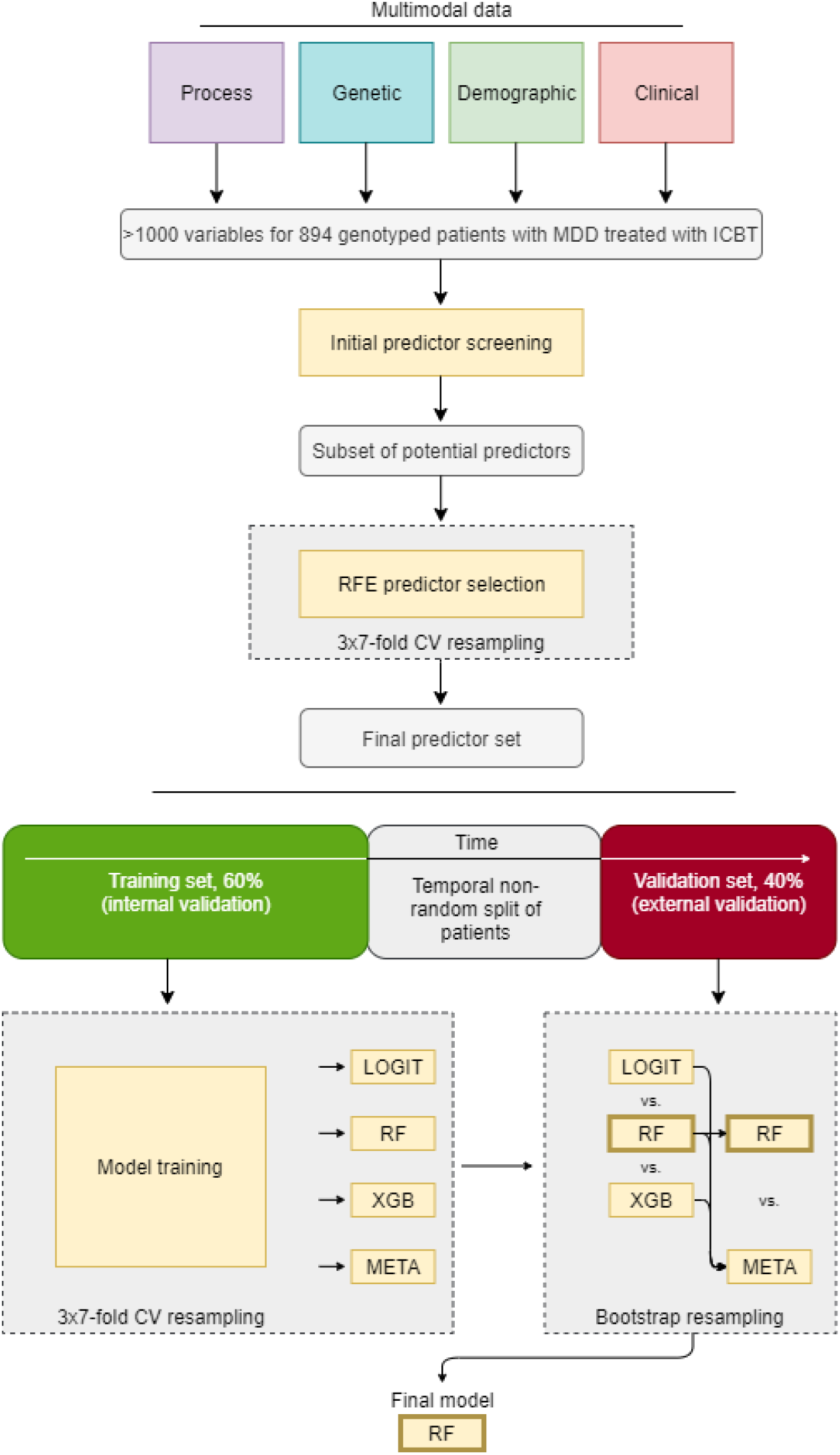
Workflow of predictor selection, model derivation, and validation. CV, cross-validation; ICBT, Internet-based Cognitive Behaviour Therapy; LOGIT, binomial logistic regression; MDD, Major Depressive Disorder; RF, random forest; RFE, recursive feature elimination; XGB, eXtreme gradient boosted trees; META, blended meta-ensemble model of LOGIT, RF, and XBG.

### Predictor selection

Only variables available at pre-treatment baseline were allowed as predictors. Genotyping and PRS calculations have been described in a previous publication.^19^ Importantly, for pure predictive modelling – and in part opposed to causal modelling – similar predictors (e.g., MADRS-S completed at screening and MADRS-S completed at pre-treatment start) were allowed as potential predictors in the same model. See^26^ and^27^ for in-depth reasoning.

Given the >1000 initial variables, psychological and psychiatric expertise (authors) was applied to screen out a subset of potential predictors. Statistical screening of predictor suitability was also applied (near-zero variance predictor cut-off: ratio 95/5 for 1^st^/2^nd^ most common categorical values or 10% unique values of total values, data missingness (>30%), and multicollinearity (r >.80). After screening, domain knowledge (authors) was again used to reduce the remaining >100 possible predictors to 69 probable predictors. To thereafter avoid human bias and achieve a robust final predictor set, algorithm selection using recursive feature elimination (RFE)^28^ was applied with 3×7-fold cross-validation resampling. At this step, the RFE served as backwards stepwise predictor selection wrapper for an inner random forest classifier using all samples (n = 894).^29^ The result defined the final predictor set used for model derivation and validation.

### Data partitioning

To further control for overfitting in addition to resampling (internal validation), data was split into two datasets based on the date of patient treatment start. Thus, a training dataset used for model development (n= 537, 60% of cases) and a hold-out validation set for model validation included only the most recent patients (n= 357, 40%). This split provides a type of external validation ^30 31^ as it also allows for temporal non-random bias to invalidate developed models.

### Model training

Models were trained using the same cross-validation as applied for feature selection. Although fairly balanced classes were present, pseudorandom downsampling of the majority class was used within each resampling fold to guarantee not overfitting to the majority class during model training and thereby also assigning equal weight to false positives and negatives. Three algorithms of deliberately increasing flexibility were used to develop predictive models on the training dataset: (i) linear main-effects logistic regression with no tuning parameters (LOGIT),^32^ (ii) Breiman-type random forest with basic grid search tuning of the tree depth hyperparameter (RF),^33^ and (iii) eXtreme Gradient Boosting machine which applies gradient descent to iteratively develop its individual trees, and here also tuned with Bayesian Optimization to better calibrate its joint optimal setting across several hyperparameters (XGB).^34^ Both (ii) and (iii) models automatically handle possible higher-order effects/interactions whereas model (i) requires manual specification of such terms and were deliberately not allowed to keep this model simple. Models (ii) and (iii) are ensemble algorithms that construct and combine a number of weak decision trees into a combined and usually more accurate model. Additionally, predicted probabilities obtained from model (i) – (iii) were also further combined into (iv) a blended meta-ensemble (META) and compared versus the top performing individual model.

### Model validation

Developed models were evaluated on the unseen hold-out validation set with retained real-world remission base rate (no downsampling) which are not perfectly balanced (44.5% in remission of total validation cases). Accuracy was the primary performance metric. We also report Area Under the receiver operating characteristics Curve (AUC, C-statistic). Associated 95% confidence bounds (CI) are provided for both outcome metrics. P-values for binomial tests of developed models versus the null model are reported, and AUC curves are also plotted against a random classifier. The null model is a no information model classifying all cases as belonging to the majority class (i.e. 55.5% correctly classified in the validation set). To test individual models against each other, their AUCs were bootstrapped (n bootstraps = 5000) and compared reporting the standard deviation of the difference between AUCs (D) and P for pairwise comparison.

### Model transparency analysis

Partial dependence plots for the two most important numeric predictors (as defined by RFE) for each of the four predictor types are provided and interpreted. Partial dependence shows the group-level influence of a single predictor on the probability for remission holding other predictors constant in the model. To further assess the influence of particular predictors on remission prediction in a single patient, the Local Interpretable Model-agnostic Explanations (LIME) procedure was applied.^35^ This modelling involves several steps, but in summary, a ridge regression model (L2 penalization) was applied with the trained RF model on permutations (n = 5000) for each individual patient and the relative influence of top 10 predictors (largest model weights) for the probability of remission in six individual patients is presented. This detailed analysis of a few individuals is not representative of all patients yet does provide more insight into how single cases are classified by the non-linear model assuming local linearity.

### Additional statistics

Continuous variables are reported as arithmetic mean (SD) and categorical variables are reported as count (%) and further specified as needed. Missing values are reported descriptively and thereafter imputed with K-Nearest Neighbor imputation using the weighted Gower distance metric and k set low (k = 3) to preserve the correlational structure of data.^36^ Analyses were performed in R^37^ using packages *AppliedPredictiveModeling, caret, caretEnsemble, corrplot, data*.*table, doParallel, dplyr, dummies, foreign, ggplot2, ggpubr, ggthemes, haven, Hmisc, lattice, latticeExtra, lime, matrixStats, mice, mlbench, MLmetrics, pdp, pROC, randomForest, rBayesianOptimization, scales, stringr, tableone, VIM, vip*, and *xgboost*.

## RESULTS

### Sample characteristics

In the full sample, observed (decimal mean [SD]; %-change [SD]; integer count (%)) pre-treatment MADRS-S total score was substantially higher (22.1 [6.3]) than at post-treatment (12.8 [5.8]) representing an almost halved (−42% [35%]) pre-post MADRS-S percentage change with 338 (43%) of patients in remission at the end of treatment.

Stratified by observed outcome, patients not in remission after treatment also scored higher (24.0 [5.8] vs 19.9 [6.0]) on pre-treatment MADRS-S and had a smaller percentage symptom decline (−22 [28] vs -68 [23]) pre-post treatment MADRS-S, compared to patients in post-treatment remission. Other outcome-stratified summary statistics are available in **Table 1 (**with full predictor explanations provided in **Table 2)** showing that patients in remission started treatment a bit earlier in the year, completed the MADRS-S questionnaire slightly earlier during the day, and completed the EQ5D questionnaire faster, compared to patients not in remission. Moreover, remitted patients were marginally younger, had a higher education and more profession-specific work experience. Patients in remission also more commonly had a diagnostic history of mild MDD, fewer depressive episodes, scored lower on MDD severity, anxiety severity, and higher on quality of life and self-efficacy, reported less alcohol and medication use (both previous and present), and were overall rated more favourably on functionality by the interviewing psychiatrist.

**Table 1.**
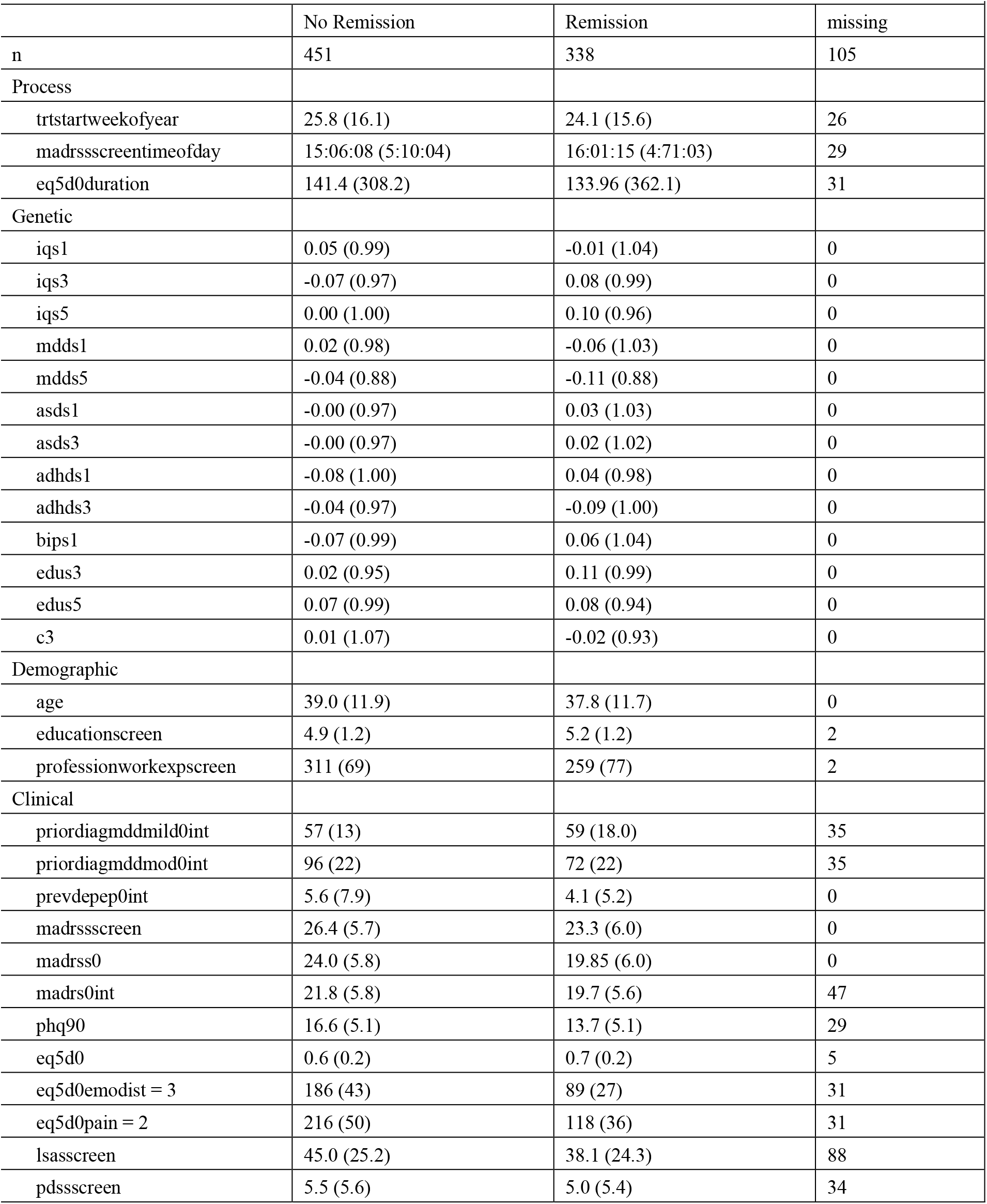
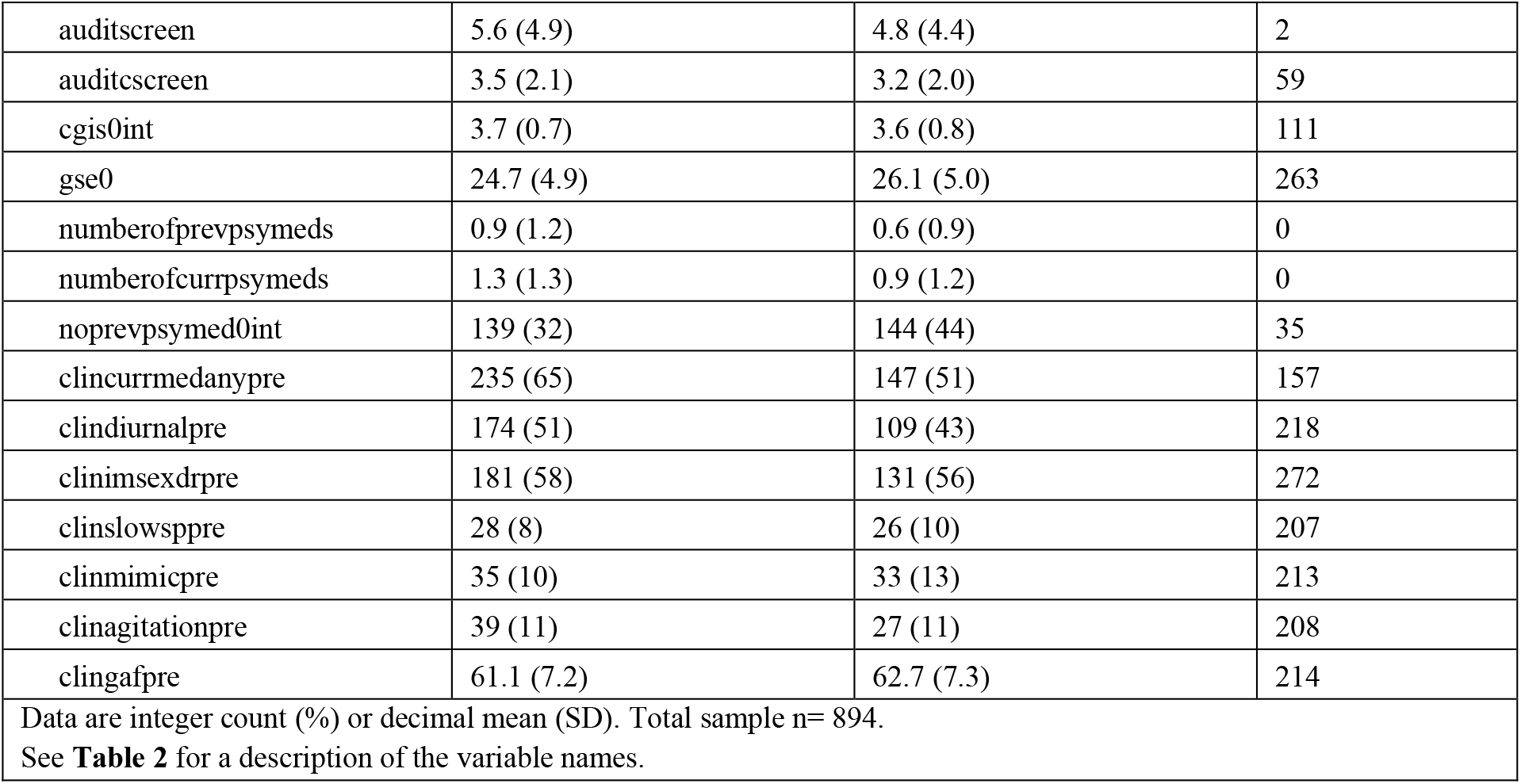
Patient summary characteristics grouped by type and stratified by outcome.

**Table 2.**
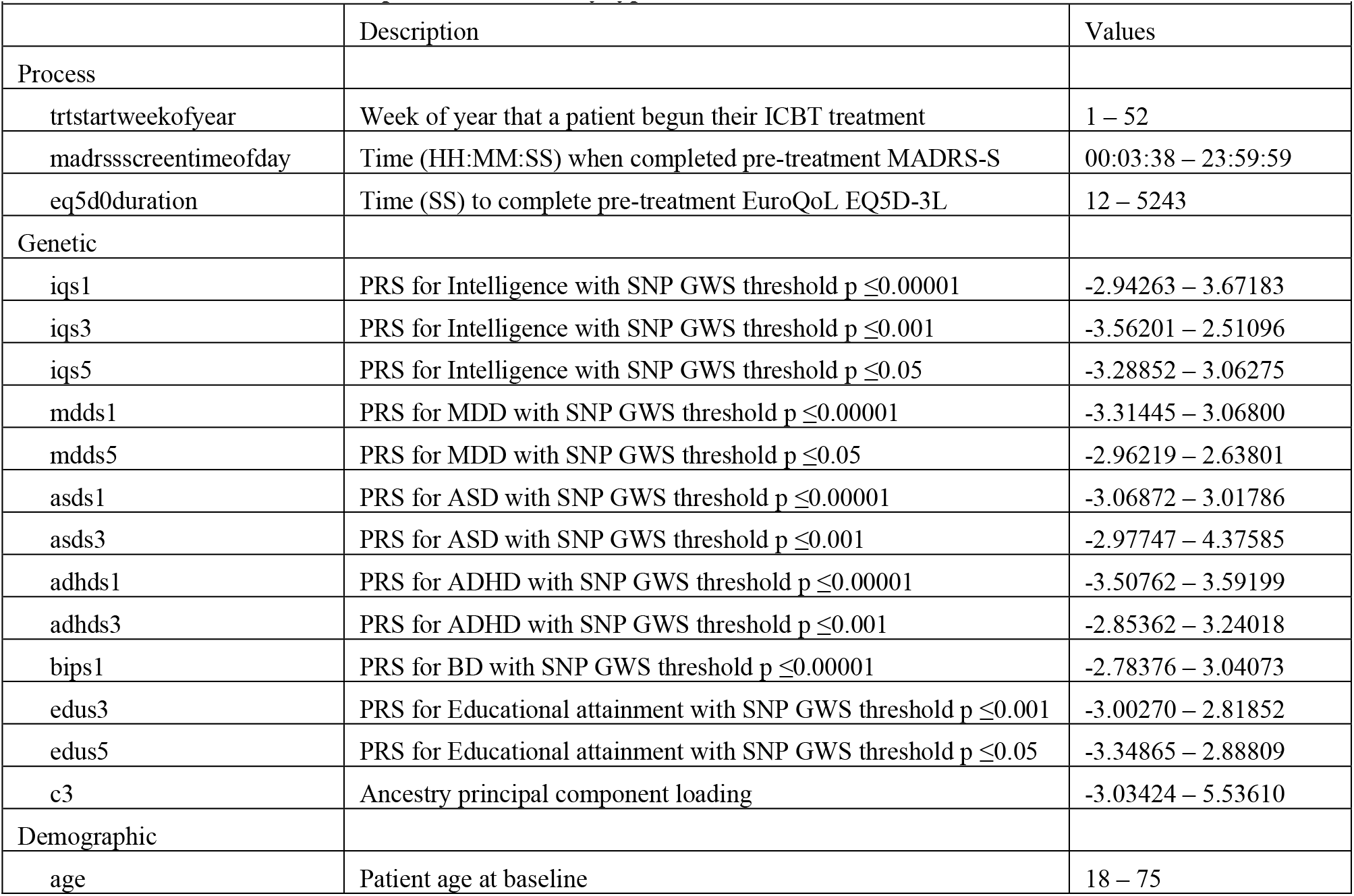
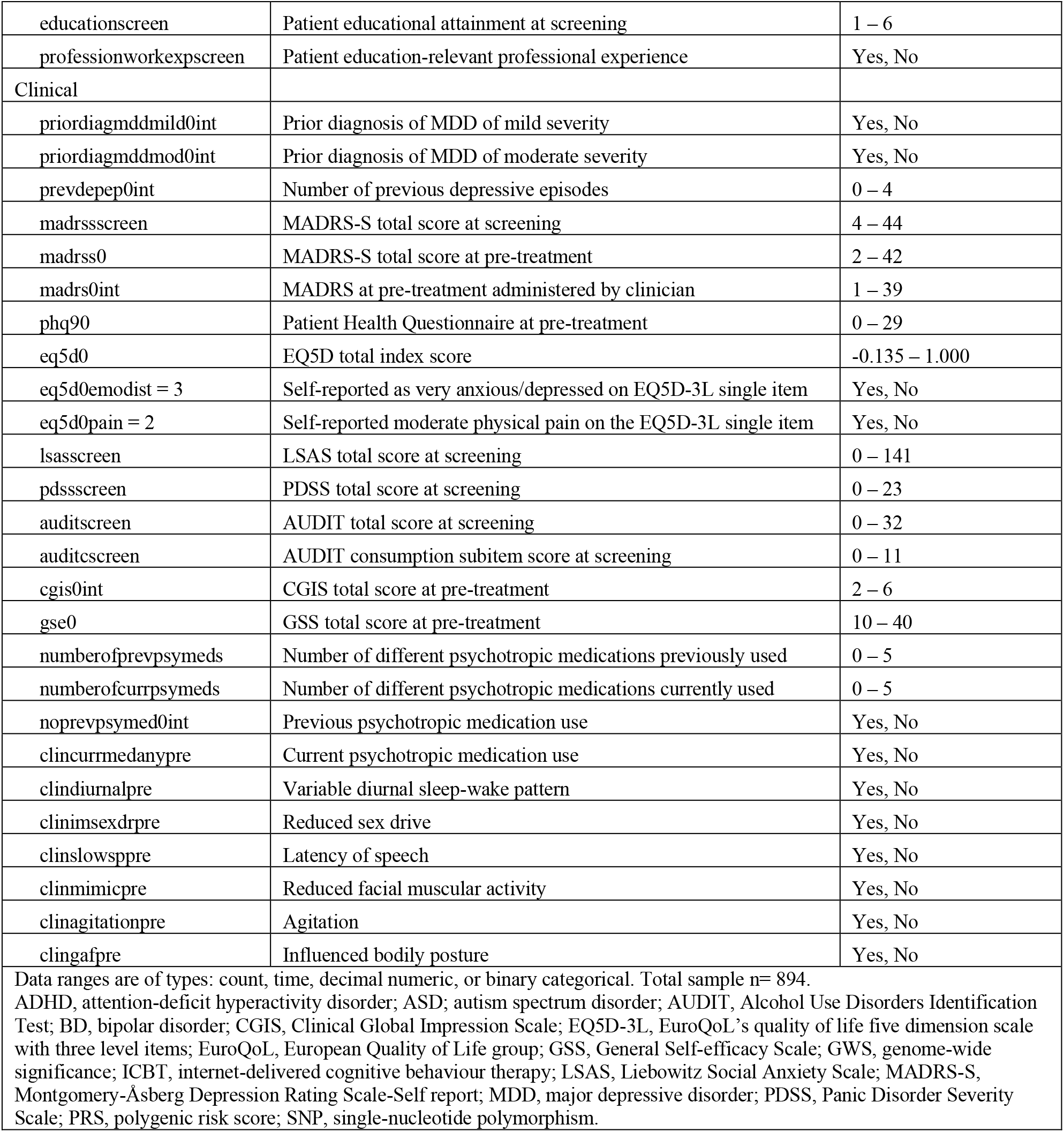
Information on the final predictors sorted by type.

### Predictor selection

Out of the 69 probable predictors, the RFE algorithm discarded 24 predictors and retained 45 predictors. This final predictor set included variables from all four predictor types (n Process= 3; Genetic= 13; Demographic= 3; Clinical= 26). **Table 2** provides further predictor details and explains the abbreviations used in the present article.

### Predictor importance

The RFE result in **Figure 2** details the individual predictors sorted by their type and their relative Gini importance (reduction in decision node impurity across trees). The most important Process predictor was time of day when the patient completed MADRS-S online, the most important Genetic predictor was the PRS for intelligence calculated with SNP GWS p ≤0.05. The most important Demographic predictor was educational attainment, and the most important Clinical predictor was pre-treatment total score MADRS-S. Overall, Clinical predictors dominated the other types with respect to both individual relative importance and total number selected by RFE. However, predictors of all types were ultimately included by the algorithm for classifying the post treatment remission target.

**Figure 2.**
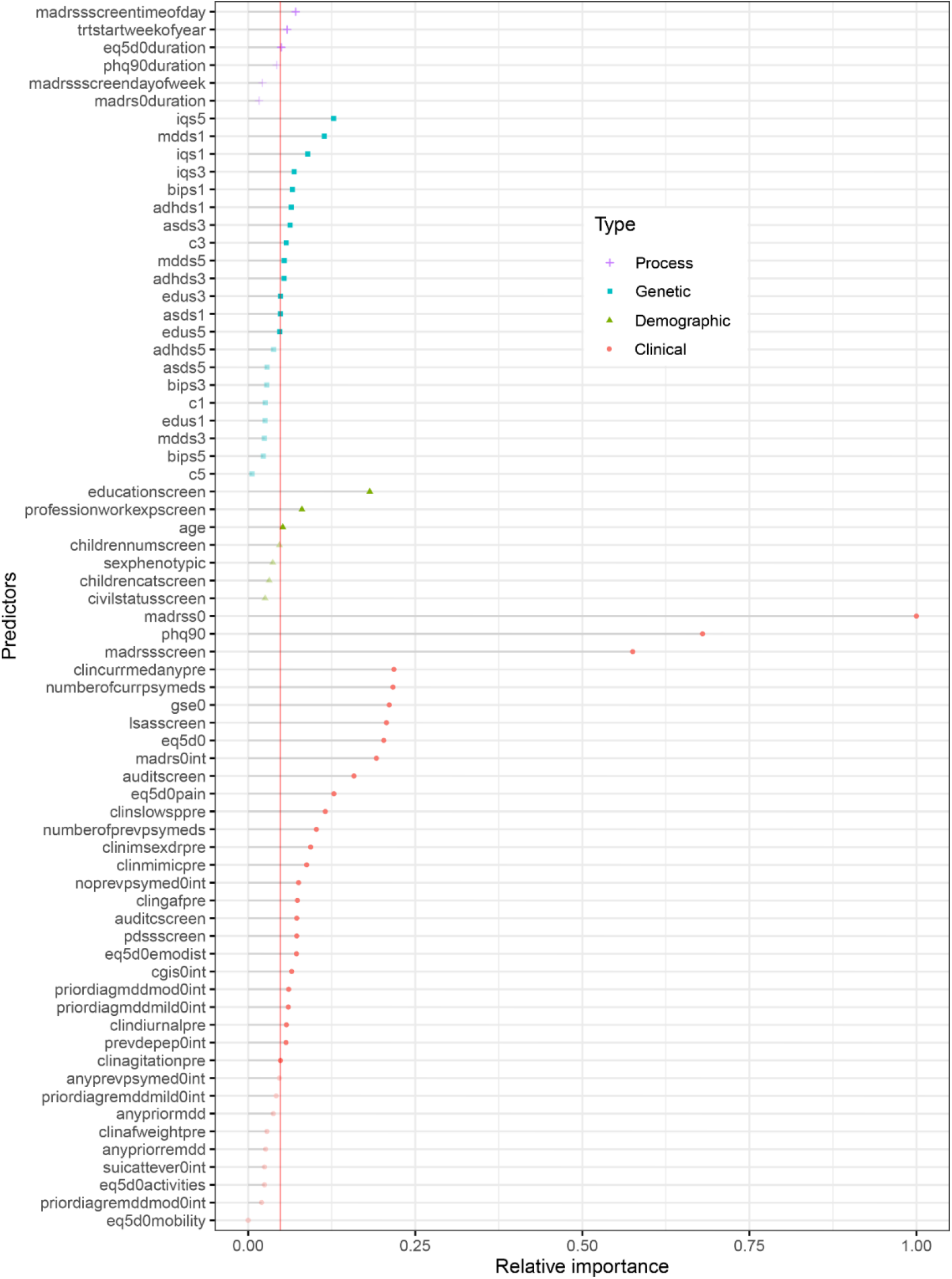
Individual predictors sorted by predictor type and relative Gini importance for predicting remission post ICBT in adults with MDD. Needle length on the x-axis represents relative importance with the strongest predictor scaled to 1 and others as proportional fractions of 1. Colour groups predictors into one of four categories (Process, Genetic, Demographic, and Clinical). The vertical line defines the RFE cut-off for final model inclusion which retained 45 variables (solid dot) and discarded 24 predictors (transparent dot). Total sample n= 894. See **Table 2** for additional predictor information. ICBT, Internet-based Cognitive Behaviour Therapy; MDD, Major Depressive Disorder; Recursive Feature Elimination

### Model validation

#### Individual model performance versus the null model

Performance was quite similar across individual models. However, RF had the best performance (Accuracy [95%CI] p vs null model) on the hold-out test set outperforming the null model (0.656 [0.604, 0.705] 0.004) which neither LOGIT (0.611 [0.558, 0.662] 0.181) nor XGB (0.613 [0.561, 0.664] 0.154) did.

#### Individual model performance versus one another

Pairing and bootstrap testing (n = 5000) of AUC curves on the hold-out validation set further showed only a statistical tendency of performance difference for LOGIT vs RF (D = 1.730, p = 0.084 favouring RF), and no significant performance difference comparing LOGIT vs XGB (D = 1.152, p = 0.249) or RF vs XGB (D = 0.463, p = 0.643). See **Figure 3** for further details. As for the Accuracy comparisons, RF had the best performance with respect to AUC.

**Figure 3.**
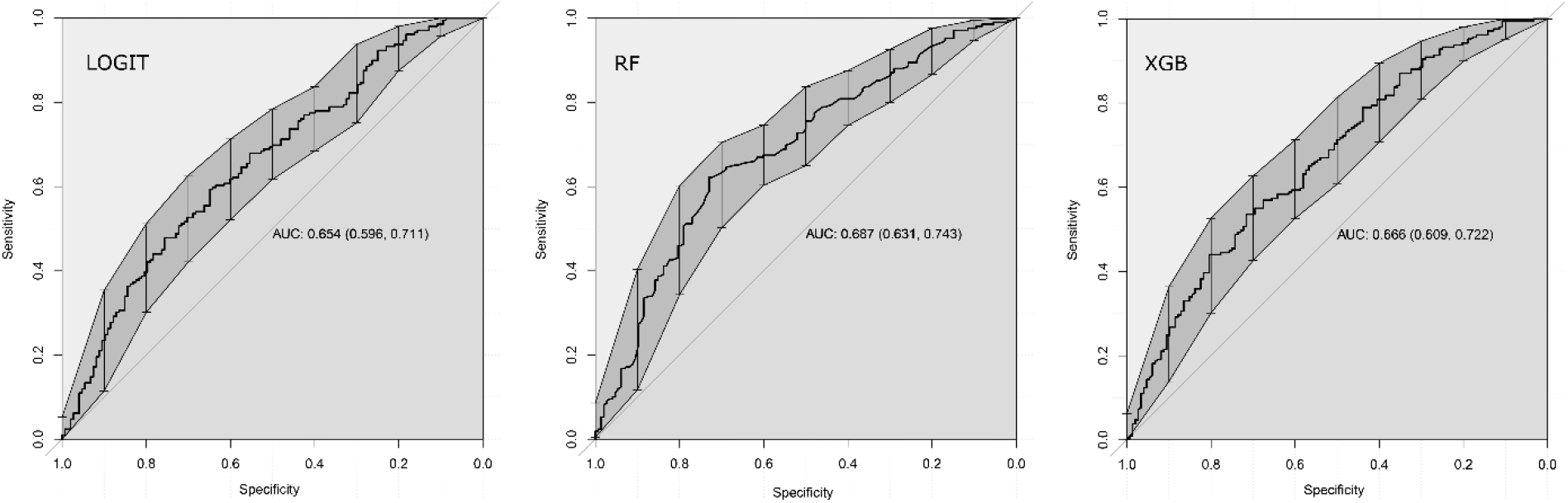
Individual AUC model performance on the hold-out test set. AUC, area under the receiver operating characteristics curve; LOGIT, logistic regression; RF, random forest; XGB, extreme gradient boosted trees

#### Meta-ensembling

Correlations of predicted probabilities across individual models were suitable for meta-ensembling (r range = 0.67 – 0.83) and a blended combination of LOGIT, RT and XGB was constructed using greedy optimization to iteratively tune individual model weights in the meta-ensemble towards local optima for each resampling fold (META). Greedy optimization can thus fail to find the global optima but is generally efficient at finding good weights across a variety of settings.^38^ Performance of META was not superior (p = 0.72) to the single RF model and RF was accepted as the final model.

#### Individual predictors

Since the best performing model was a non-linear ensemble classifier, straightforward interpretation of single predictor coefficients was impossible. Instead, partial dependence of most important numeric predictors sorted in pairs by predictor type are available in **Figure 4** showing individual predictor contributions to the probability of remission after ICBT when all other variables are mean centred and held constant in the RF model. For Process predictors, we found that completing MADRS-S screening in the internet portal later in the day and starting treatment earlier in the year predicted remission. Regarding Genetics, having an intermediate or low PRS for depression predicted non-remission whereas a higher PRS for intelligence predicted remission. There was also some additive predictive power for remission having both a high PRS for depression and intelligence. With respect to Demographics, a lower age and higher education predicted remission. Considering Clinical predictors, having a low score on pre-treatment PHQ-9 and MADRS-S additively predicted remission.

**Figure 4.**
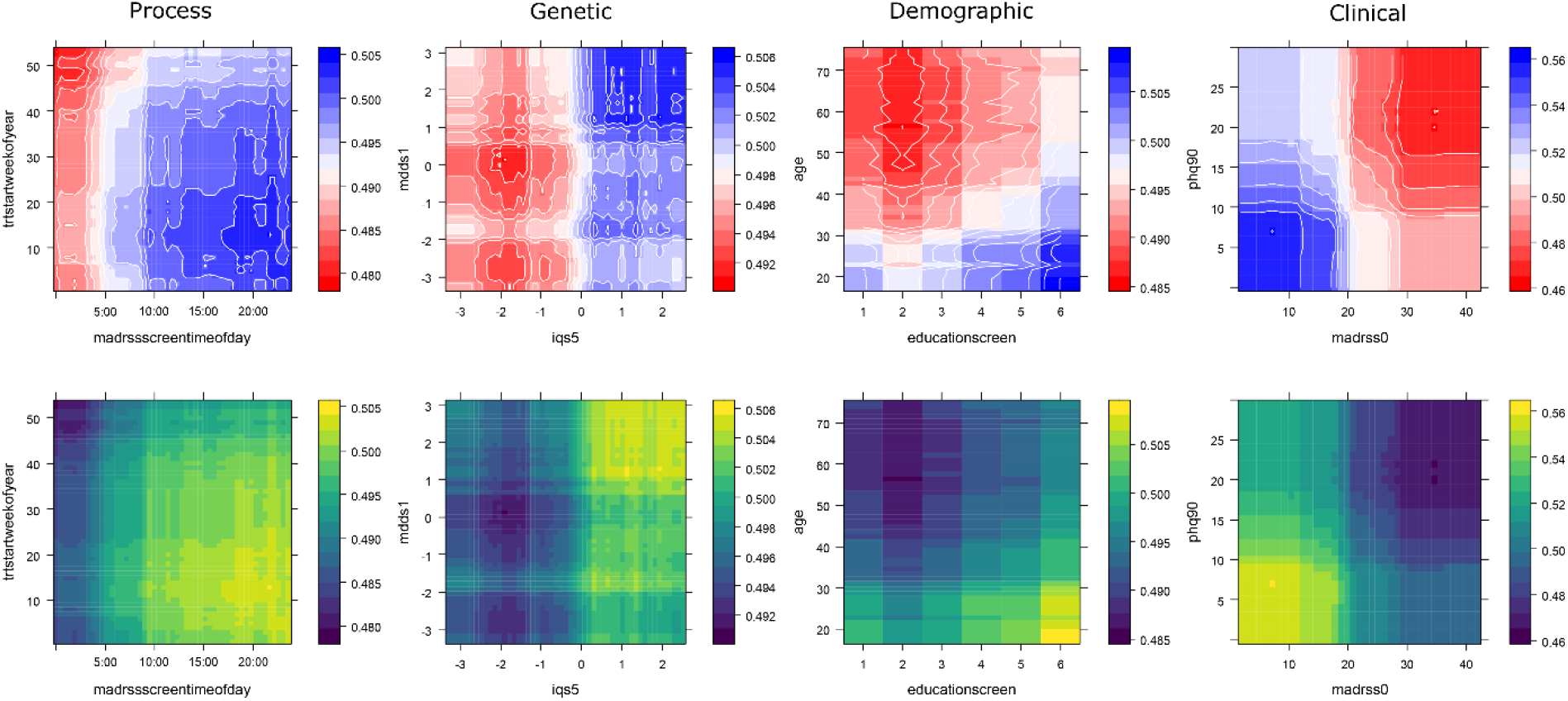
Partial prediction plots of strongest bivariate predictors by predictor type in the final random forest model. Each column represents one of the four predictor types from which the two top numeric predictors have been plotted on their respective x and y axis. Colour represents the individual variable contribution to the predicted probability of remission in two ways. The upper row is colour-blindness friendly (Blue = higher probability of remission) and the bottom row is greyscale compatible (Light = higher probability of remission). The probability contribution of each variable in each panel is calculated when all other predictors in the model are centred and held constant. See **Table 2** for additional predictor information.

#### Individual patient predictions

How predictors influenced the probability for post ICBT remission in six individual patients is available in **Figure 5** as LIME plots showing that the RF classifier reused several of the strongest predictors (e.g. MADRS-S, PRS for intelligence, latency of speech, PHQ-9) across patients. For example, a score below 19 indicated remission (Patient 3, Patient 6) but a score above 26 on MADRS-S contradicted remission (Patient 1, 2) and the pattern was similar for the other symptom scales. Also, a higher genetic PRS for intelligence indicated remission (Patient 2, 3) and lower contradicted remission (Patient 1, 4, 5, 6), a GSE score above 29 indicated remission (Patient 4). Having education-relevant professional experience indicated remission (Patient 2, 3, 5, 6) and vice versa (Patient 1, 4). Overall, scoring higher than 5 on AUDIT (alcohol), reporting substantial QoL-related physical pain, and taking more than one psychiatric medicine contradicted remission, whereas having a high education indicated remission. Finally, the model would classify Remission in Patient 2, 3, 6 and No Remission in Patient 1, 4, and 5.

**Figure 5.**
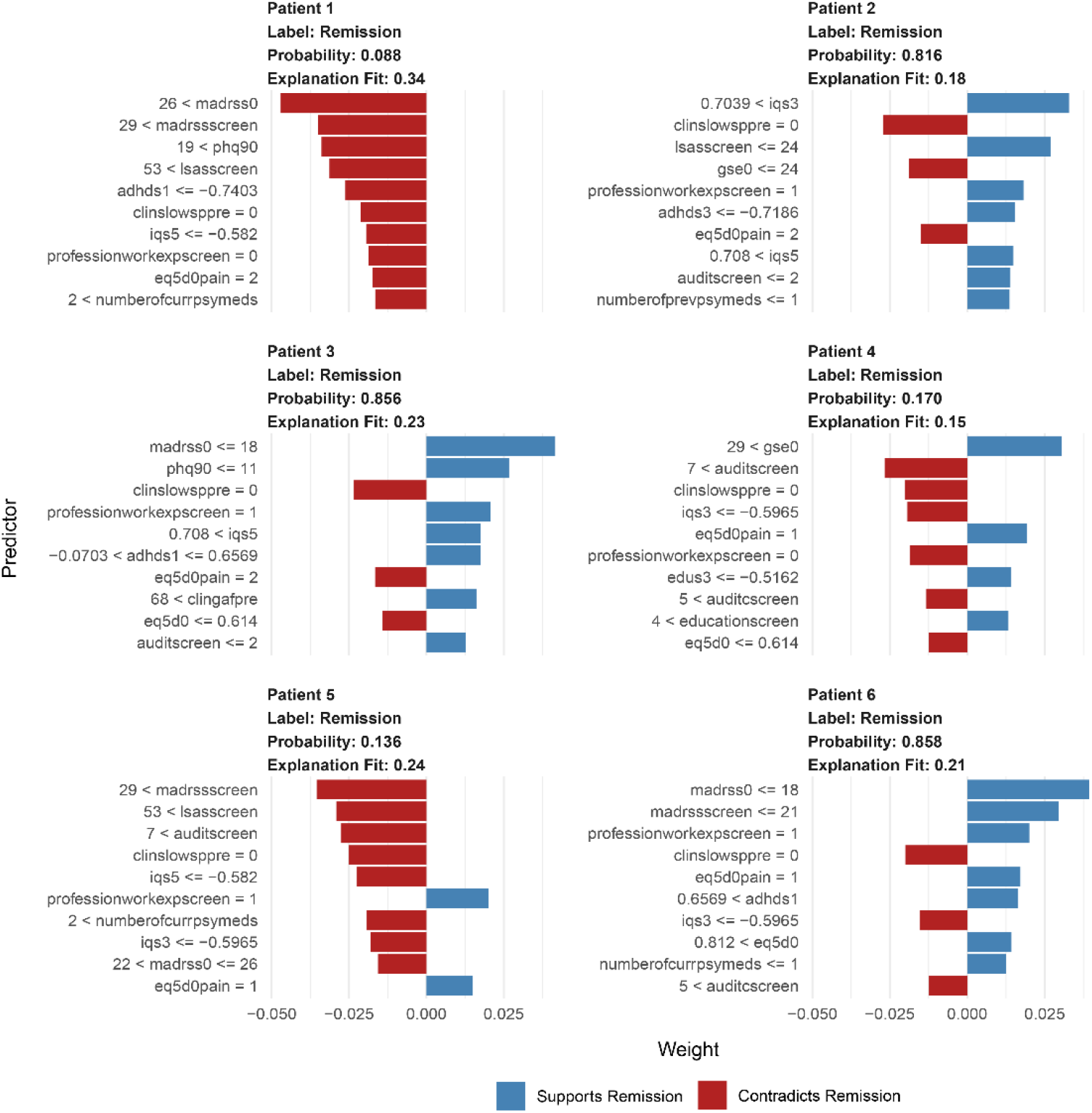
Individual case predictions by the RF model exemplified with six individual patients. The top 10 predictors and their cut-off values influencing the probability for post ICBT remission in each patient. Data are from the final RF model with additional modelling using the LIME framework. See **Table 2** for additional predictor information.

## DISCUSSION

The present study investigated a multi-modal prediction approach within a contemporary machine learning pipeline for predicting remission in a large sample of adult patients with MDD treated with ICBT in routine psychiatric care in Sweden. Both at the group and individual patient level, the final model made use of all four of the investigated predictor types (Process, Genetic, Demographic, and Clinical), thus confirming some predictors but also expanding on previous psychiatric predictor-finding studies. Although clinical predictors were strongest, PRSs for intelligence, depression, and other traits, and also ICBT process-specific predictors, and demographic predictors, independently contributed to predicting remission. Modelling was (a) deliberately incrementally increased in complexity from log-linear main effects regression to non-linear and fine-tuned gradient boosting and meta-ensembling, (b) internally validated through robust resampling and (c) externally validated on unseen, temporally separated data. Models were fairly accurate but with remaining room for improvement, suggesting a potential for future multi-modal prediction and implementation of such models for routine care prediction of post ICBT remission in patients with MDD.

Our findings corroborate previous findings of symptom severity, education, age, medication use, and PRSs as predictors for ICBT outcomes.^3 8 10 12 13 16 19^ Regarding remission of MDD in particular, our result is in line with Andersson and colleagues^11^ in that depression symptom severity and higher anxiety predicted the outcome. Interestingly, the number of predictors that our final model settled with was fairly high (n = 45), also consistent with the logic of a multi-modal approach for predicting complex traits and behaviours. Considering genetics, we only partially replicated the findings from a prior study^19^ to the extent that a signal for the PRS for autism was found, although it was quite weak and surpassed by both the PRS for intelligence and PRS for MDD. The previous study used the same patients but it investigated symptom change over time using a linear mixed model, whereas the present study employed the use of multimodal data and machine learning for non-linear binary classification of remission post treatment. One can speculate that a genetic propensity for MDD would likely fit the treatment well, and as this to a substantial degree does translate into higher MDD symptoms in phenotype, such patients will numerically have both a greater room for improvement and be disproportionately subject to regression to the mean. Further, the present finding of PRS for intelligence seems reasonable given that ICBT is a highly verbal treatment, posing demands on the patient to not only be computer- and internet-proficient but also able to express fairly abstract concepts such as their internal emotions through an abstract medium (written text), and also to decide, plan, execute behavioural change. Having a high genetic propensity for intelligence in such a situation is likely beneficial for treatment success which would then feed into the probability of remission.

The present study employed the classic approach of baseline pre-to-post prognostication. Previous studies have used symptom change during early parts of ICBT to predict final outcome^17 39^ and a recent RCT suggests that such individual patient predictions can be of actual clinical benefit.^40^ A main strength of within-treatment prediction is a probabilistic basis for adapting treatment while it is ongoing.^17^ The strength of baseline prediction does however offset a weakness-by-design in within-treatment prediction, i.e. that baseline prediction provides the probabilistic basis for matching correct treatment to a patient before it is even initiated and thereby omitting the risk of termination, wasted resources, and unnecessary patient suffering. Future evidence may favour prediction from either baseline for guiding treatment matching or prediction within-treatment for adaptation. It is however likely that both approaches will be useful in a future, more data-driven psychiatry and today we view them as complementary.

### Clinical implications

The present verification of known Clinical and Demographic predictors strengthens these predictors and their potential place in routine care for adults with MDD undergoing ICBT. The identification of online Process predictors, as well as Genetic predictors suggests future possible pathways towards improved remission prediction in these patients. There are however important additional challenges when progressing from (a) identifying new group-level predictors for ICBT outcomes in MDD to (b) individual patient-level predictions that matter in clinical reality. Herein, we demonstrate how (a) → (b) modelling could be performed for individual patients. More research is however needed to ascertain the extent by which these predictions could provide triage decision support for groups of patients and/or guide treatment choice for the individual patient (e.g. predicted low probability of being in remission after ICBT could be offered alternative treatment before starting ICBT (face-to-face CBT, home-based CBT, medication).

The prediction performance of the best model herein was good although with room for improvement before clinical implementation is pursued. The next step would therefore be to extend the present model with a larger sample, and also more predictors. Another extension would be to treat unaggregated SNPs as a high-dimensional input space to a deep learning model, a class of algorithms known to perform particularly well with high-dimensional data,^41^ because aggregate PRSs, as used herein, might result in the loss of potentially useful genetic information. Given the complex nature of MDD and remission after ICBT, phenotype registry predictors – which are uniquely available in national Swedish registries – such as birth complications and detailed medical history could also contribute additional predictive power. Another ambitious follow-up study would be a prediction model test with adults with MDD in Sweden, preferably as a superiority trial with randomization of clinicians to the prediction model as decision support for initial treatment choices versus treatment as usual. Effectiveness and cost-effectiveness could then be evaluated on both remission immediately after completed treatment and also on long-term outcomes of medication and healthcare utilisation, rehospitalisation, accidents, and suicide. The nationwide IPSY clinic with its high volume of patients undergoing ICBT, standardized protocol and online procedure with registered process data would be the natural setting for such a trial. The design could also be extended to other patient groups and ICBT treatment sites. A third important future study which demands relatively minor resources would be a test of the present model predictions against experienced clinicians predicting the same outcome as the model in a prospective sample of patients. That design could ascertain model prediction accuracy directly relative to human benchmark performance.

### Strengths and limitations

A recent review^42^ on precision psychiatry highlighted some general shortcomings, including that only 15% of developed prediction models were validated and, out of those, 94% remained under high risk of other potential biases. Naturally, more flexible models are prone to overfitting, yet the present sample size allowed for both thorough internal resampling validation and also temporally separate hold-out data for external model validation. The aforementioned review also found that unimodal prediction model studies reported better performance than multimodal counterparts, questioning the potential benefit of the latter.^42^ Our findings do not support that notion, and neither do other findings showing that MDD is a polygenic trait influenced by a multitude of factors.^14 15^ Remission in MDD is in turn a complex behavioural outcome that is, for instance, causally influenced by evidence-based psychotherapy. Although a predictor is not necessarily predicated on causality, one may for complex multi-factorial phenomena expect a plethora of causal agents, and consequently, several diverse predictors. Instead, it could be that models in the field in general have suboptimal quality, that sample sizes in precision psychiatry tend to be relatively small coupled with the curse of dimensionality, and that there is an underutilisation of more flexible algorithms better able to detect subtly predictive patterns in complex data than linear main effect models are. Regarding generalisability, the present model was developed with data from routine specialised care in Sweden and may not immediately generalise to other clinics, beyond our national border, or to other patient groups. This is however also a strength because the high ecological validity means that results should be readily implementable at the nationwide recruiting IPSY clinic for patient treated with ICBT for MDD. Models often need to be retrained and weights adjusted to fit new contexts which could be accomplished given that samples are available from other clinics in Sweden or abroad. Patient online behaviour is logged automatically in every comprehensively designed ICBT platform, and from the log database one can extract and calculate Process variables that are rarely used as predictors for ICBT outcomes. This is potentially an underutilised source of predictive power for ICBT outcomes models and more research on this class of predictors seems warranted.^12 43^

## CONCLUSION

A new, multi-modal machine learning model for predicting MDD remission status after treatment with ICBT in routine care was derived and empirically validated. In future iterations, this model may inform tailored intervention before the start of ICBT for MDD. The multi-modal approach to predicting remission and similar complex traits in MDD patients was supported and warrants further investigation.

## Data Availability

Data includes sensitive personal information and is not available freely by default. Data can be made available to external researcher(s) given reasonable request and following proper procedure, including study ethical approval and adhering to the limits of patient informed consent.

## Acknowledgements

We are deeply grateful to the patients and their clinicians at the IPSY clinic.

## Author contributions

JW, JB, and CR designed the study. JW analysed data and drafted the manuscript. All authors interpreted the findings, critically revised the manuscript, and approved its final form and submission. All authors agree to be accountable for all aspects of the work.

